# Biomarker-Informed Phenotyping of the Heart Failure Continuum: A Population-Based Evaluation of the Second Universal Definition

**DOI:** 10.64898/2026.11.04.26367312

**Authors:** Haoxin Chen, Jiancheng Ye

## Abstract

**Objective:** To develop and evaluate a biomarker-informed, computable phenotype of the heart failure (HF) continuum, aligned with 2026 AHA/ACC/ESC/WHF Expert Consensus, using NHANES and linked mortality data, and to determine whether this phenotype identifies subgroups with distinct mortality risks.

**Methods:** We analyzed 11,477 U.S. adults aged ≥20 years from the 1999–2004 NHANES cycles who participated in the surplus-sera biomarker substudy and were linked to National Death Index follow-up. Participants were classified into four phenotypes according to the 2026 Universal Definition of Heart Failure: no apparent risk, Stage A (at risk), Stage B (elevated NT-proBNP without self-reported HF), and Stage C (self-reported HF). Survey-weighted Kaplan–Meier curves and nested, multiply imputed Cox proportional hazards models, with sequential adjustment for demographic, socioeconomic, and cardiometabolic covariates, were used to estimate associations with mortality. Pairwise phenotype comparisons, standardized adjusted survival estimates, and restricted mean survival times were also calculated. Incremental prognostic value was evaluated by comparing discrimination and reclassification metrics between the full phenotype and a conventional three-level model that excluded NT-proBNP.

**Results:** Weighted prevalences were 16.9%, 70.0%, 10.7%, and 2.4% for no apparent risk, Stage A, Stage B, and Stage C, respectively. In fully adjusted models, the hazard ratios (HRs) relative to no apparent risk were 1.58 (95% CI, 1.08–2.30), 2.88 (95% CI, 2.00–4.16), and 2.56 (95% CI, 1.61–4.09), respectively. Stage B was associated with significantly higher mortality risk than Stage A (HR, 1.83; 95% CI, 1.62–2.07), whereas the difference between Stage C and Stage B was not statistically significant (HR, 0.89; 95% CI, 0.71–1.11). The association for Stage B attenuated less across sequentially adjusted models than that for Stage C. Adjusted 10-year mortality risks were 15.2% for Stage B and 14.2% for Stage C. Incorporating Stage B into the conventional model yielded only marginal improvements in discrimination (ΔC-statistic, 0.002; 95% CI, 0.001– 0.002).

**Conclusion:** A biomarker-informed phenotype of the HF continuum can be operationalized using national survey data and identifies a pre-HF subgroup with an adjusted mortality risk not significantly different from that of individuals with overt HF, despite only marginal gains in prognostic discrimination.

## INTRODUCTION

Heart failure (HF) affects an estimated 6.7 million U.S. adults, and its prevalence is projected to increase.[1, 2] For decades, the American College of Cardiology/American Heart Association (ACC/AHA) have conceptualized HF as a continuum encompassing asymptomatic individuals at risk (Stage A), structural or functional cardiac abnormalities without symptoms (Stage B), symptomatic HF (Stage C), and advanced disease (Stage D).[3–5] An estimated one-third of adults meet Stage A criteria, while 24–34% may meet Stage B criteria.[1] Community-based cohorts have demonstrated that asymptomatic Stage A/B disease is associated with substantial risk of progression,[6] yet identifying these stages remains challenging outside specialized research settings.[7]

The concept of Stage B originated in the 2001 ACC/AHA guideline,[3] which defined this stage by the presence of structural heart disease in the absence of HF symptoms. The 2021 First Universal Definition of HF renamed this stage “pre-HF” and expanded its diagnostic criteria to include elevated natriuretic peptides or cardiac troponins as corroborating evidence.[8] The 2022 AHA/ACC/HFSA guideline subsequently incorporated this framework into U.S. clinical practice, recognizing that individuals with HF risk factors and elevated BNP or NT-proBNP, or persistently elevated cardiac troponin, may meet Stage B criteria even in the absence of imaging-confirmed structural abnormalities.[5, 8] An analysis from the Atherosclerosis Risk in Communities (ARIC) study demonstrated that biomarker-based criteria reclassified approximately one in five older adults from Stage A to Stage B, identifying individuals with higher subsequent risk.[9] The 2026 Second Universal Definition reaffirmed this diagnostic pathway, while introducing greater international harmonization, designating Stage 0 for individuals without HF risk factors (corresponding to the “no apparent risk” group in this study; Table S1), and emphasizing Stage B as an opportunity for early detection.[10]

Because natriuretic peptide concentrations increase with age independently of cardiac disease, the choice of diagnostic threshold, particularly fixed versus age-adjusted cutoffs, can materially influence pre-HF classification.[11] This raises the possibility that pre-HF could be operationalized using biomarkers alone, extending its identification beyond imaging-based cohorts. However, whether explicitly incorporating biomarker criteria into a staged phenotype, rather than treating NT-proBNP solely as a continuous risk marker, meaningfully differentiates prognosis at the population level remains unclear. Most HF epidemiologic studies rely on self-reported diagnoses, thereby primarily capturing Stage C disease, whereas natriuretic peptide studies generally evaluate these biomarkers as continuous predictors rather than components of a staged phenotype. Although one large study demonstrated that NT-proBNP independently predicts mortality outside a formal staging framework,[12] related NHANES research examining preclinical HF, defined by conventional risk factors and prior myocardial infarction, and all-cause and cardiovascular mortality over 1999–2018 did not establish a biomarker-defined Stage B or evaluate the full HF continuum.[13] Moreover, elevated biomarkers in the absence of conventional risk factors may still indicate clinically meaningful risk but may not be adequately represented by any single conventional stage.[14] Accordingly, whether biomarker-defined Stage B constitutes a prognostically distinct phenotype relative to both no apparent risk and established HF remains unresolved.[15]

NHANES provides a valuable opportunity to address this question. NT-proBNP was measured in stored surplus serum samples from NHANES participants examined during 1999–2004,[16] and these survey cycles also collected self-reported HF status in a nationally representative sample of U.S. adults, with prospective mortality follow-up through linkage to the National Death Index. Previous analyses of this biomarker substudy demonstrated that NT-proBNP was a robust predictor of mortality independent of kidney function, [17] and documented racial and ethnic differences in NT-proBNP concentrations,[18] highlighting both the feasibility and the complexities of its application in population-based research. Collectively, these data provide an opportunity to construct a four-level phenotype comprising no apparent risk, Stage A, biomarker-defined Stage B, and self-reported Stage C, and to preliminarily evaluate its prognostic relevance.

This study aims to develop a computable, biomarker-informed phenotype of the HF continuum, aligned with consensus criteria, using NHANES and linked mortality data, and to determine whether formal staging provides prognostic information beyond a general elevation in mortality risk. We hypothesized that the 2026 Universal Definition could be operationalized using variables available in a nationally representative health survey and that the resulting phenotypes would exhibit distinct long-term mortality trajectories, providing preliminary population-level evidence relevant to the updated staging framework.

## METHODOLOGY

### Study design

We conducted a cohort analysis of participants from the 1999–2000, 2001–2002, and 2003–2004 NHANES cycles. NHANES is a nationally representative, multistage survey of the noninstitutionalized U.S. population conducted by the National Center for Health Statistics. The HF-continuum phenotype was assigned cross-sectionally at baseline, and participants were subsequently followed for mortality. These survey cycles were selected because NT-proBNP measurements were available only during this period, in a subset of participants selected for the NHANES surplus-sera biomarker substudy. Mortality outcomes were ascertained through linkage to the National Death Index, with follow-up time calculated from the examination date to death or administrative censoring on December 31, 2019.

The analytic framework translated the 2026 Universal Definition into a four-level phenotype: no apparent HF risk, Stage A (at risk), Stage B (elevated NT-proBNP without self-reported HF), and Stage C (self-reported HF). Figure 1 illustrates the mapping of each consensus construct to its corresponding NHANES operational definition and indicates whether it could be measured directly, approximated using proxy variables, or not meaningfully assessed. Because several constructs could not be operationalized with the available data, the analysis was restricted to the four phenotypes that could be constructed and evaluated in relation to mortality.

**Figure 1.**
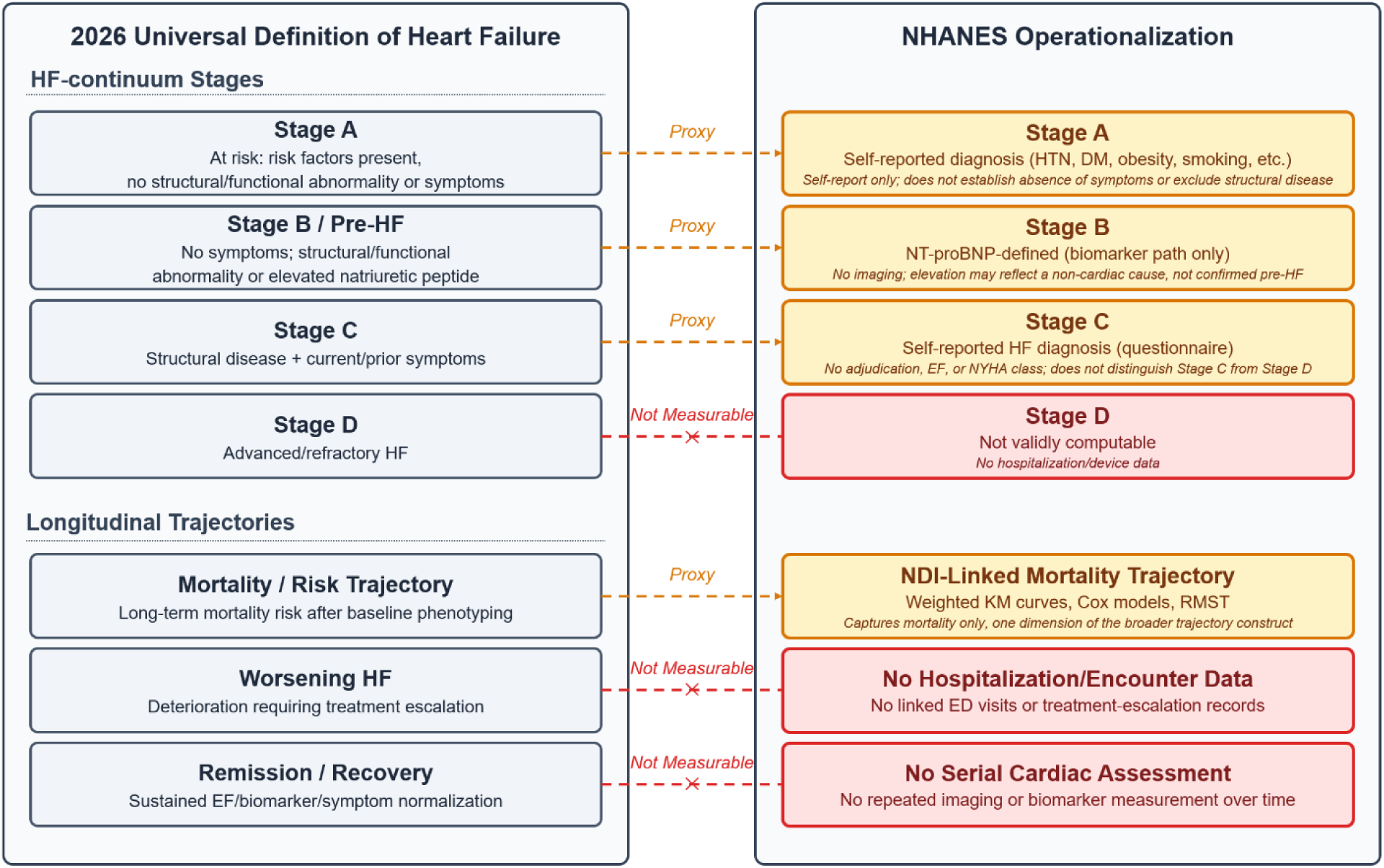
Conceptual framework.

Participants were eligible if they were aged ≥20 years at examination and had nonmissing self-reported HF status, complete examination and survey design variables, eligibility for mortality linkage, recorded follow-up time, and nonmissing core phenotype variables. Pregnant participants were excluded. All analyses accounted for the complex survey design, including primary sampling units and strata. The pooled 6-year biomarker substudy weight was derived by combining two-thirds of the 1999–2002 4-year weight with one-third of the 2003–2004 2-year weight, using the appropriate weight variable for each participant’s survey cycle. Variance estimation was performed using Taylor series linearization implemented through the survey package in R. For discrimination and reclassification metrics not amenable to linearization, variance was estimated using cluster bootstrap resampling.

### Stage B/pre-HF biomarker

Stage B was defined using NT-proBNP concentrations measured in stored serum samples from the NHANES surplus-sera biomarker substudy. NT-proBNP is renally cleared, and its concentrations may be lower in individuals with obesity independently of cardiac status. Moreover, a single fixed threshold does not account for the well-established age-related increase in NT-proBNP concentrations. Consistent with the general principles of the Universal Definition and threshold approaches adopted in previous studies,[19] participants were classified as Stage B if their NT-proBNP concentration exceeded the 90th percentile of an age-and sex-specific reference distribution. This distribution was constructed using an unweighted approach across 14 strata defined by seven 10-year age groups and sex, among participants with nonmissing NT-proBNP measurements and no self-reported HF. Percentile ranks were calculated within each stratum, with tied values assigned identical percentile ranks. Strata containing fewer than 10 reference participants were considered insufficient for percentile estimation. Accordingly, Stage B status was classified as missing for participants with missing NT-proBNP measurements, missing age, or membership in an undersized stratum. These participants were excluded from the analysis unless they met the criteria for Stage C. The prevalence of Stage B in the overall cohort was not expected to equal exactly 10%, because the reference distribution was constructed without survey weighting, whereas cohort prevalence estimates incorporated the complex survey design, and because Stage B classification was restricted to participants without Stage C.

### Phenotype assignment and descriptive analysis

The four-level phenotype was assigned hierarchically, prioritizing Stage C over Stage B, Stage B over Stage A, and Stage A over no apparent risk (corresponding to Stage 0 in the 2026 framework). Each participant was assigned to a single mutually exclusive category, with lower-priority stages assigned only after higher-priority criteria had been positively ruled out, rather than by default when classification at a higher level was not possible. Stage C was defined by self-reported physician-diagnosed congestive HF on the questionnaire. Among participants not meeting Stage B or Stage C criteria, Stage A was defined by the presence of at least one of 10 risk factors. Eight factors—hypertension, diabetes, obesity, chronic kidney disease (CKD), albuminuria, prior coronary heart disease (CHD) or myocardial infarction (MI), prior stroke, and current smoking— corresponded to risk factors specified in the 2026 Universal Definition. Age ≥65 years[20] and a composite social vulnerability indicator [21] were additionally included based on prior literature. The latter was defined by the presence of at least one of the following: low income, low educational attainment, lack of health insurance, or absence of a usual source of healthcare. Because social vulnerability was included among the 10 Stage A criteria, the no apparent risk group was consequently more advantaged with respect to income, education, and insurance coverage.[22]

Phenotype distributions were summarized using unweighted counts and survey-weighted percentages with 95% confidence intervals (CIs). Baseline characteristics were compared across phenotypes using survey-weighted percentages for categorical variables and survey-weighted means for continuous variables, each with corresponding 95% CIs. Survey-weighted phenotype prevalence was further examined across age groups, sex, race and ethnicity, poverty-income ratio, CKD status, and diabetes status.

### Survival analysis

The primary outcome was all-cause mortality, ascertained through linkage to the National Death Index (NDI), with follow-up extending from the examination date to death or censoring. Unadjusted survival across phenotypes was estimated using survey-weighted Kaplan–Meier methods. Adjusted associations were evaluated using survey-weighted Cox proportional hazards models in a nested sequence. Model 0 was unadjusted and characterized the overall association. Model 1 adjusted for age, sex, and race and ethnicity. Model 2 additionally adjusted for educational attainment, poverty-income ratio, and health insurance status. Model 3, the primary model, further adjusted for hypertension, diabetes, obesity, CKD, smoking, and prior CHD, MI, or stroke. NT-proBNP was not included as a covariate in any model because it was used to define Stage B and would therefore be structurally collinear with the exposure. Pairwise comparisons among Stages A, B, and C were performed by changing the reference category and refitting Model 3.

Adjusted 10-year mortality risks were estimated through marginal standardization. Specifically, 10-year survival probabilities were predicted for each participant under each phenotype, with all other covariates held at their observed values, and then averaged using survey weights. These estimates represent predicted risks under hypothetical phenotype reassignment across the cohort’s covariate distribution, rather than observed risks among participants currently classified within each phenotype or causal effects of changing phenotype.

Missing covariate data were addressed using multiple imputation by chained equations, generating 20 imputed datasets. Mortality status, follow-up time, and phenotype assignment were included as fully observed predictors but were not imputed. Cox models were fitted separately within each imputed dataset, and estimates were pooled using Rubin’s rules. Adjusted survival probabilities, restricted mean survival time (RMST), and differences in RMST relative to the no-apparent-risk group were derived from the primary model. Incremental prognostic value was assessed by comparing a conventional three-level phenotype without NT-proBNP against the full four-level phenotype using the C-statistic, calibration slope, integrated discrimination improvement (IDI), and continuous net reclassification improvement (NRI).

Sensitivity analyses evaluated the robustness of the findings to alternative phenotype definitions and analytical specifications, including substitution of a fixed NT-proBNP threshold for the percentile-based definition, exclusion of participants with advanced CKD based on measured estimated glomerular filtration rate (eGFR), stratification by obesity status, complete-case analysis, use of age as the underlying time scale in Cox models, and exclusion of deaths occurring within six months of examination.

## RESULTS

### Analytic cohort construction

Among 31,126 participants in NHANES 1999–2004, 15,332 were aged ≥20 years, of whom 15,262 had nonmissing self-reported HF status. Because NT-proBNP was measured only in the subset selected for the surplus-sera biomarker substudy, and valid subsample-specific survey weights were required to obtain nationally representative estimates, participants without a valid substudy weight were excluded (n = 2,998), leaving 12,264 participants. Further exclusions were made for ineligibility for mortality linkage (n = 12), incomplete core phenotype variables (n = 125), and pregnancy at the time of examination (n = 650), resulting in a final analytic cohort of 11,477 participants (Figure 2). Over a median follow-up of 16.8 years (interquartile range [IQR], 14.8–18.6), 3,483 deaths were recorded.

**Figure 2.**
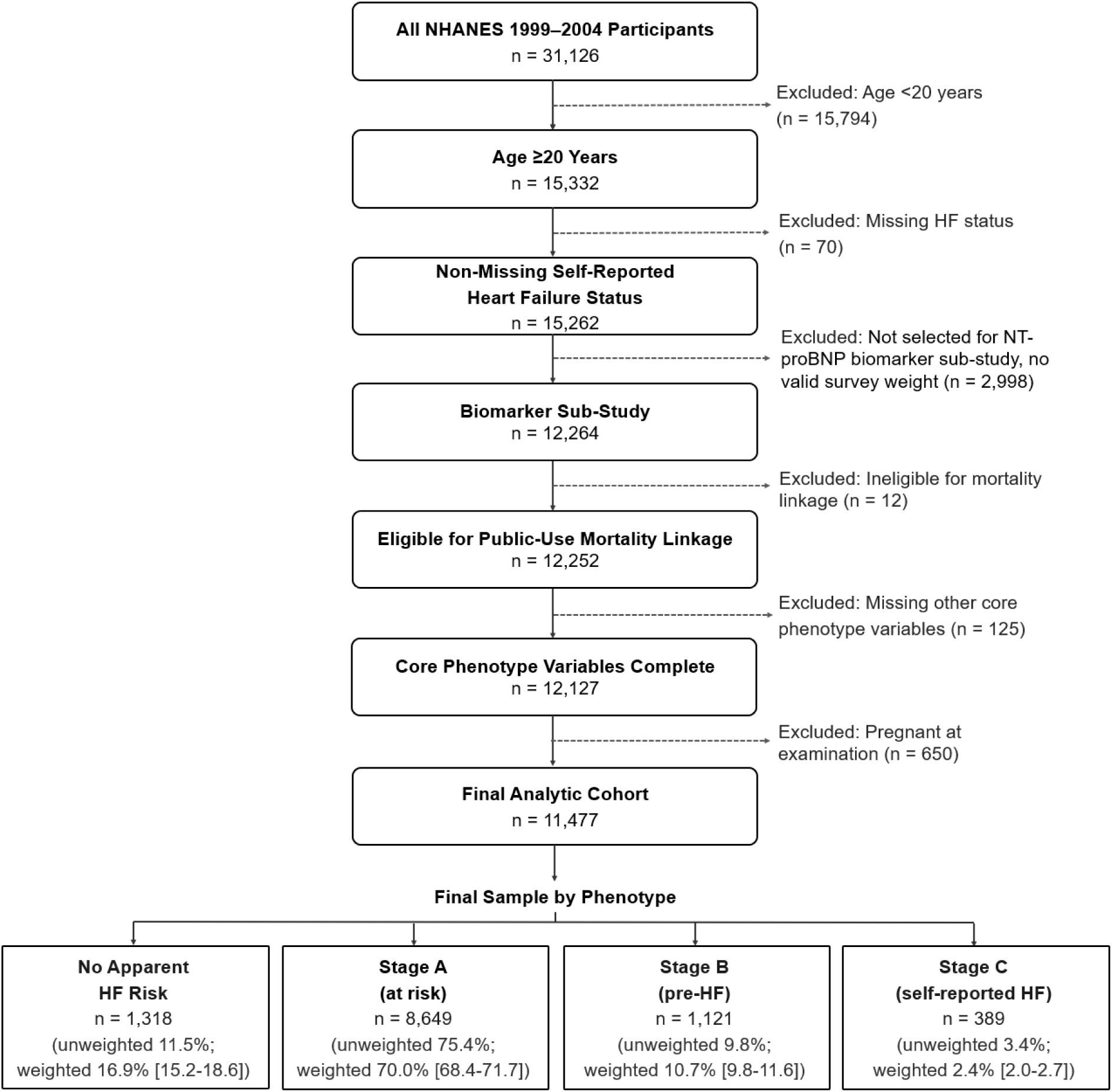
Cohort construction diagram.

### Distribution of HF phenotypes

Under the four-level HF classification, 1,318 participants were classified as having no apparent HF risk, 8,649 met Stage A criteria, 1,121 met Stage B criteria (elevated NT-proBNP without self-reported HF), and 389 were classified as Stage C based on self-reported HF. The corresponding survey-weighted prevalences were 16.9% (95% CI, 15.2–18.6), 70.0% (95% CI, 68.4–71.7), 10.7% (95% CI, 9.8–11.6), and 2.4% (95% CI, 2.0–2.7), respectively.

Survey-weighted phenotype distributions varied across demographic and clinical subgroups (Figure 3). No apparent risk and Stage A exhibited the most pronounced age-related gradients: the prevalence of no apparent risk was negligible among individuals aged ≥70 years, whereas Stage C prevalence increased to 11.3% among those aged ≥80 years. Phenotype distributions were broadly similar between sexes. Non-Hispanic White participants and those classified in the other-race category had higher proportions of no apparent risk and Stage B, and a lower proportion of Stage A, than the remaining racial and ethnic groups, which exhibited broadly comparable distributions. Stage C prevalence was highest among participants living below the poverty threshold (4.0%) and lowest among those with a poverty-income ratio (PIR) >4 (1.2%), whereas the distribution of no apparent risk showed the opposite pattern. Stage A prevalence varied modestly by CKD status (65.1% among participants with CKD vs. 70.2% among those without CKD) and diabetes status (76.3% among those with diabetes vs. 69.4% among those without diabetes). In contrast, Stage B and Stage C were substantially more prevalent among participants with CKD or diabetes, with more pronounced differences by CKD status than by diabetes status. Because PIR, CKD, and diabetes were incorporated into the Stage A definition, subgroup comparisons involving these variables are subject to inherent definitional overlap.

**Figure 3.**
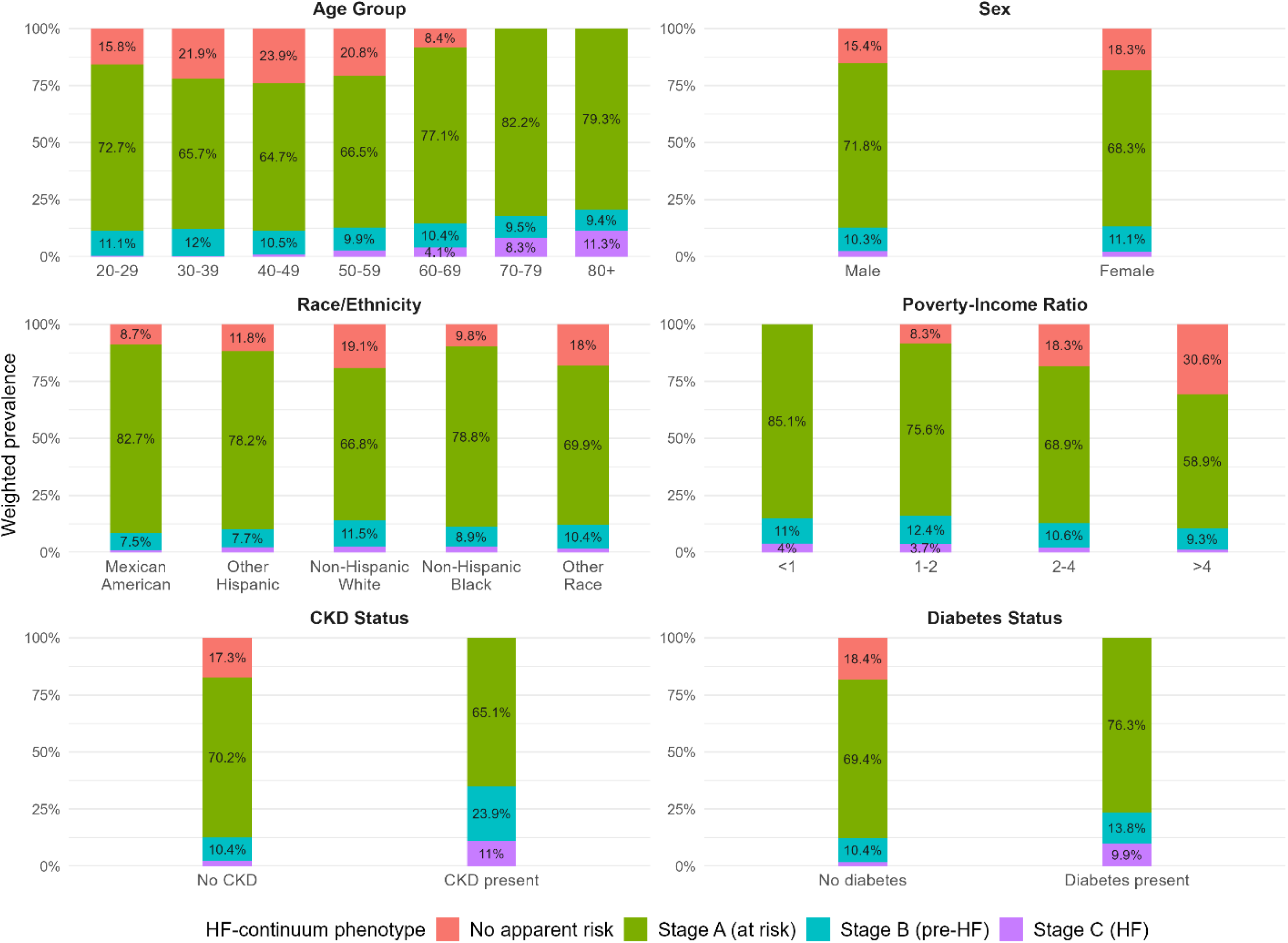
Weighted prevalence of HF phenotypes by demographics and clinical strata.

### Baseline characteristics across phenotypes

Baseline characteristics differed substantially across the four phenotypes (Table 1), with the most pronounced differences observed for variables incorporated into the Stage A and Stage B definitions.

**Table 1.** Baseline characteristics by HF phenotype.

| Variable | No apparent HF risk | Stage A (at risk) | Stage B (pre-HF) | Stage C (self-reported HF) | p-value |
|---|---|---|---|---|---|
| Age, weighted mean (95% CI), years | 41.4 (40.6–42.1) | 47.2 (46.6–47.9) | 46.0 (45.1–47.0) | 65.6 (63.4–67.8) | <0.001 |
| Female, No. (weighted % [95% CI]) | 726 (55.7% [53.4–58.1]) | 4218 (50.1% [49.1–51.2]) | 555 (53.1% [49.6–56.6]) | 174 (48.8% [41.7–55.9]) | 0.003 |
| Race/ethnicity, No. (weighted % [95% CI]) |  |  |  |  | <0.001 |
| Mexican American | 165 (3.7% [2.6–4.9]) | 2147 (8.5% [6.5–10.5]) | 189 (5.0% [3.0–7.1]) | 67 (3.3% [1.6–4.9]) |  |
| Other Hispanic | 47 (4.0% [2.1–5.9]) | 419 (6.4% [4.0–8.9]) | 39 (4.2% [2.1–6.2]) | 14 (5.6% [0.3–10.8]) |  |
| Non-Hispanic White | 877 (81.3% [78.1–84.4]) | 4144 (68.5% [65.0–71.9]) | 661 (77.5% [73.2–81.7]) | 238 (77.0% [70.8–83.2]) |  |
| Non-Hispanic Black | 171 (6.3% [4.9–7.7]) | 1660 (12.2% [10.0–14.5]) | 193 (9.1% [6.7–11.4]) | 61 (11.1% [6.4–15.8]) |  |
| Other Race/ Multi-Racial | 58 (4.7% [3.1–6.3]) | 279 (4.4% [3.4–5.4]) | 39 (4.3% [2.8–5.8]) | 9 (3.1% [0.5–5.7]) |  |
| Education, No. (weighted % [95% CI]) |  |  |  |  | <0.001 |
| Less than 9th grade | 0 (0.0% [0.0–0.0]) | 1529 (8.2% [7.4–9.0]) | 182 (7.7% [6.0–9.3]) | 115 (20.3% [15.6–24.9]) |  |
| 9-11th grade | 0 (0.0% [0.0–0.0]) | 1628 (16.4% [15.2–17.5]) | 204 (15.2% [12.7–17.8]) | 89 (21.9% [16.5–27.2]) |  |
| High school grad/GED | 288 (20.3% [17.8–22.9]) | 2100 (27.7% [25.9–29.4]) | 284 (27.0% [24.3–29.6]) | 78 (24.7% [18.7–30.7]) |  |
| Some college/AA degree | 457 (32.9% [29.3–36.5]) | 2104 (29.2% [27.8–30.6]) | 258 (26.8% [23.5–30.1]) | 76 (23.6% [17.9–29.3]) |  |
| College graduate+ | 573 (46.8% [42.5–51.1]) | 1267 (18.6% [17.0–20.3]) | 193 (23.3% [19.1–27.6]) | 30 (9.6% [5.4–13.9]) |  |
| Poverty-Income Ratio, weighted mean (95% CI) | 4.0 (3.9–4.1) | 2.8 (2.7–2.9) | 2.8 (2.7–2.9) | 2.2 (2.0–2.4) | <0.001 |
| Uninsured, No. (weighted % [95% CI]) | 0 (0.0% [0.0–0.0]) | 1985 (22.3% [20.8–23.8]) | 198 (18.2% [15.3–21.2]) | 24 (8.1% [3.6–12.5]) | <0.001 |
| Usual Source of Care, No. (weighted % [95% CI]) |  |  |  |  | <0.001 |
| Has a usual place | 1301 (98.8% [98.1–99.6]) | 7006 (79.8% [78.4–81.2]) | 971 (87.2% [84.1–90.3]) | 370 (94.2% [91.2–97.3]) |  |
| More than one place | 17 (1.2% [0.4–1.9]) | 66 (0.7% [0.5–1.0]) | 15 (1.2% [0.4–2.0]) | 8 (2.1% [0.8–3.4]) |  |
| No usual place | 0 (0.0% [0.0–0.0]) | 1574 (19.5% [18.1–20.8]) | 135 (11.6% [9.0–14.2]) | 11 (3.7% [1.1–6.3]) |  |
| Household Food Security, No. (weighted % [95% CI]) |  |  |  |  | <0.001 |
| Full food security | 1175 (94.9% [93.4–96.5]) | 6497 (82.4% [81.0–83.8]) | 851 (81.9% [79.1–84.8]) | 280 (77.0% [70.6–83.4]) |  |
| Marginal food security | 38 (2.3% [1.3–3.3]) | 631 (6.0% [5.1–6.9]) | 73 (5.7% [4.0–7.4]) | 42 (11.0% [6.2–15.7]) |  |
| Low food security | 29 (1.9% [1.2–2.7]) | 789 (7.1% [6.3–7.9]) | 94 (7.3% [5.4–9.1]) | 36 (7.7% [4.3–11.1]) |  |
| Very low food security | 13 (0.9% [0.2–1.5]) | 408 (4.5% [3.8–5.1]) | 64 (5.1% [3.7–6.5]) | 16 (4.3% [1.6–7.0]) |  |
| Hypertension, No. (weighted % [95% CI]) | 0 (0.0% [0.0–0.0]) | 3038 (32.4% [30.6–34.2]) | 483 (36.3% [32.9–39.8]) | 275 (65.9% [60.0–71.8]) | <0.001 |
| Systolic BP, weighted mean (95% CI), mmHg | 115.8 (114.8–116.8) | 125.1 (124.5–125.7) | 126.4 (124.7–128.0) | 131.9 (129.2–134.7) | <0.001 |
| Diastolic BP, weighted mean (95% CI), mmHg | 72.5 (71.8–73.3) | 72.0 (71.5–72.6) | 71.5 (70.7–72.4) | 68.4 (66.0–70.8) | 0.003 |
| Diabetes, No. (weighted % [95% CI]) | 0 (0.0% [0.0–0.0]) | 908 (7.8% [7.1–8.5]) | 146 (9.3% [7.1–11.5]) | 129 (29.8% [23.8–35.7]) | <0.001 |
| HbA1c, weighted mean (95% CI), % | 5.2 (5.1–5.2) | 5.5 (5.5–5.6) | 5.4 (5.4–5.5) | 6.0 (5.9–6.1) | <0.001 |
| BMI, weighted mean (95% CI), kg/m2 | 24.7 (24.5–25.0) | 29.0 (28.7–29.2) | 27.7 (27.1–28.3) | 30.4 (29.3–31.5) | <0.001 |
| Waist Circumference, weighted mean (95% CI), cm | 87.5 (86.9–88.2) | 98.5 (98.0–99.0) | 95.5 (93.9–97.0) | 104.6 (102.6–106.6) | <0.001 |
| Dyslipidemia, No. (weighted % [95% CI]) | 528 (48.1% [45.2–51.0]) | 5134 (73.0% [71.8–74.2]) | 638 (66.7% [63.0–70.5]) | 291 (79.5% [74.9–84.1]) | <0.001 |
| Never Smoked >=100 Cigarettes, No. (weighted % [95% CI]) | 953 (70.8% [67.7–73.9]) | 4140 (46.2% [44.1–48.2]) | 472 (41.5% [37.9–45.1]) | 145 (35.7% [29.6–41.8]) | <0.001 |
| Current Smoking, No. (weighted % [95% CI]) | 0 (0.0% [0.0–0.0]) | 2204 (30.4% [28.7–32.0]) | 329 (31.7% [28.1–35.3]) | 70 (21.2% [15.6–26.8]) | <0.001 |
| Chronic Kidney Disease, No. (weighted % [95% CI]) | 0 (0.0% [0.0–0.0]) | 212 (2.0% [1.7–2.4]) | 78 (4.9% [3.8–6.0]) | 45 (10.2% [6.8–13.6]) | <0.001 |
| Albuminuria, No. (weighted % [95% CI]) | 0 (0.0% [0.0–0.0]) | 1288 (13.0% [12.0–13.9]) | 298 (19.4% [16.6–22.2]) | 119 (26.3% [21.0–31.6]) | <0.001 |
| eGFR, weighted mean (95% CI), mL/min/1.73m2 | 102.2 (100.8–103.7) | 99.7 (98.8–100.6) | 95.0 (93.2–96.8) | 74.2 (70.2–78.2) | <0.001 |
| Urine Albumin-Creatinine Ratio, weighted mean (95% CI), mg/g | 6.3 (5.9–6.6) | 22.8 (19.7–26.0) | 92.4 (60.8–124.0) | 163.6 (72.1–255.1) | <0.001 |
| CKD Risk Category, No. (weighted % [95% CI]) |  |  |  |  | <0.001 |
| Very high risk | 2 (0.1% [0.0–0.3]) | 107 (0.9% [0.6–1.1]) | 90 (4.3% [3.1–5.5]) | 56 (10.6% [7.1–14.1]) |  |
| High risk | 0 (0.0% [0.0–0.0]) | 388 (3.2% [2.8–3.5]) | 125 (8.1% [6.5–9.7]) | 57 (14.3% [10.1–18.5]) |  |
| Moderately increased risk | 15 (1.3% [0.8–1.9]) | 1108 (10.5% [9.7–11.3]) | 171 (12.3% [9.9–14.6]) | 112 (25.4% [20.6–30.2]) |  |
| Low risk | 1301 (98.5% [98.0–99.1]) | 7046 (85.5% [84.6–86.4]) | 735 (75.3% [72.5–78.2]) | 164 (49.7% [43.9–55.5]) |  |
| Coronary Heart Disease / MI, No. (weighted % [95% CI]) | 0 (0.0% [0.0–0.0]) | 441 (4.1% [3.5–4.6]) | 153 (10.7% [8.7–12.7]) | 232 (60.3% [55.1–65.6]) | <0.001 |
| Stroke, No. (weighted % [95% CI]) | 0 (0.0% [0.0–0.0]) | 278 (2.5% [2.1–2.9]) | 63 (3.7% [2.5–5.0]) | 70 (16.5% [11.5–21.5]) | <0.001 |
| NT-proBNP, weighted arithmetic mean (95% CI), pg/mL | 46.5 (44.1–48.9) | 74.2 (70.9–77.5) | 530.5 (459.4–601.7) | 1063.9 (838.7–1289.2) | <0.001 |
| NT-proBNP, weighted geometric mean (95% CI), pg/mL | 33.9 (31.9–36.0) | 39.6 (38.2–41.0) | 224.1 (209.8–239.2) | 302.3 (247.0–370.0) | <0.001 |
| Log NT-proBNP, weighted mean (95% CI) | 3.5 (3.5–3.6) | 3.7 (3.6–3.7) | 5.4 (5.3–5.5) | 5.7 (5.5–5.9) | <0.001 |
| Self-Rated Health, No. (weighted % [95% CI]) |  |  |  |  | <0.001 |
| Excellent | 523 (40.5% [37.4–43.5]) | 1396 (18.0% [16.9–19.1]) | 160 (16.5% [13.8–19.2]) | 10 (2.2% [0.6–3.7]) |  |
| Very good | 500 (39.7% [37.0–42.5]) | 2233 (30.3% [28.8–31.8]) | 256 (28.2% [24.7–31.8]) | 38 (9.1% [5.7–12.5]) |  |
| Good | 238 (16.2% [13.9–18.5]) | 2968 (33.7% [32.4–35.0]) | 360 (31.8% [28.1–35.4]) | 102 (28.6% [22.0–35.3]) |  |
| Fair | 49 (2.9% [2.1–3.8]) | 1676 (14.8% [13.8–15.9]) | 259 (18.2% [15.2–21.2]) | 147 (34.4% [28.2–40.7]) |  |
| Poor | 8 (0.7% [0.1–1.2]) | 368 (3.1% [2.5–3.8]) | 84 (5.3% [3.6–7.1]) | 91 (25.7% [19.6–31.8]) |  |

Note: Dyslipidemia was defined as physician-reported high cholesterol, total cholesterol ≥ 240 mg/dL, or HDL cholesterol <40 mg/dL (men) or <50 mg/dL (women). Negative lower confidence bounds were truncated at 0.0%.

#### Demographic and socioeconomic characteristics

Mean age increased across the HF continuum, reaching 65.6 years in Stage C, whereas sex distributions were broadly similar across phenotypes. Racial and ethnic composition differed most notably in Stage A, which was predominantly non-Hispanic White (68.5%), consistent with this largest phenotype more closely reflecting the underlying population distribution. Educational attainment and socioeconomic status declined progressively across phenotypes: the proportion of college graduates decreased from 46.8% in the no-apparent-risk group to 9.6% in Stage C, while mean PIR declined from 4.0 to 2.2. Uninsured individuals accounted for 8.1% of Stage C. Fewer than 20% of participants overall reported lacking a usual source of healthcare, and full food security was most prevalent in the no-apparent-risk group (94.9%).

#### Cardiometabolic risk factors

Hypertension, absent by definition in the no-apparent-risk group, increased in prevalence to 65.9% in Stage C. Systolic blood pressure increased, whereas diastolic blood pressure declined across progressively advanced phenotypes, consistent with age-related widening of pulse pressure. Diabetes prevalence was comparable between Stages A and B but increased to 29.8% in Stage C, accompanied by higher HbA1c levels. BMI, waist circumference, and dyslipidemia were also highest in Stage C and lowest in the no-apparent-risk group. Current smoking was most prevalent in Stages A and B but less common in Stage C despite greater cumulative tobacco exposure, potentially reflecting smoking cessation following clinical diagnosis.

#### Kidney function and renal markers

The prevalence of self-reported CKD and albuminuria, as well as KDIGO very-high-risk classification, increased progressively across phenotypes. Mean eGFR declined from 102.2 mL/min/1.73 m² in the no-apparent-risk group to 74.2 mL/min/1.73 m² in Stage C. Conversely, the urine albumin-to-creatinine ratio increased markedly by Stage B (92.4 mg/g) and remained elevated in Stage C (163.6 mg/g), suggesting that renal abnormalities were already prominent at the pre-HF stage.

#### Ischemic and cerebrovascular history

Prior CHD/MI and stroke were absent by definition in the no-apparent-risk group and were most prevalent in Stage C, affecting 60.3% and 16.5% of participants, respectively.

#### Biomarker profile

NT-proBNP concentrations increased substantially across phenotypes, with weighted geometric means of 33.9 pg/mL in the no-apparent-risk group, 39.6 pg/mL in Stage A, 224.1 pg/mL in Stage B, and 302.3 pg/mL in Stage C.

#### Self-rated health

Self-rated health deteriorated progressively across phenotypes. The proportion reporting excellent health decreased from 40.5% in the no-apparent-risk group to 2.2% in Stage C, in which fair or poor health ratings collectively predominated.

### Mortality risks and trajectories across phenotypes

Over the follow-up period, 58, 2,645, 463, and 317 deaths occurred in the no-apparent-risk, Stage A, Stage B, and Stage C groups, respectively, corresponding to 23,177.1, 132,691.5, 15,330.4, and 3,465.2 person-years of observation (Table 2). Survey-weighted mortality rates increased progressively across phenotypes, from 2.16 per 1,000 person-years in the no-apparent-risk group to 13.92, 18.89, and 73.18 per 1,000 person-years in Stages A, B, and C, respectively.

**Table 2.** Mortality risks by HF phenotype.

|  | **No apparent risk** | **Stage A** | **Stage B** | **Stage C** |
|---|---|---|---|---|
| Deaths, No. (unweighted) | 58 | 2645 | 463 | 317 |
| Person-Years (unweighted) | 23177.1 | 132691.5 | 15330.4 | 3465.2 |
| Mortality Rate per 1,000 Person-Years, weighted (95% CI) | 2.16 (1.47–2.86) | 13.92 (13.21–14.62) | 18.89 (17.06–20.72) | 73.18 (62.14–84.23) |
| Adjusted 10-Year Mortality Risk, % | 7.0 (5.6–8.4) | 10.0 (9.5–10.4) | 15.2 (14.3–16.2) | 14.2 (12.8–15.7) |
| Adjusted RMST Difference at 10 Years, mo | Reference | −1.66 (−2.49 - −0.86) | −4.84 (−5.68 - −3.98) | −4.21 (−5.53 - −2.83) |

Note: Death counts and person-years are unweighted. Mortality rates are survey-weighted incidence rates per 1,000 person-years, with 95% confidence intervals (CIs) accounting for the complex survey design. Adjusted 10-year mortality risks and restricted mean survival time (RMST) differences were estimated by standardizing predictions from the fully adjusted model (Model 3) to the analytic cohort, with 95% CIs obtained using cluster bootstrap resampling (200 replicates). RMST differences are expressed relative to the no-apparent-risk group.

Survey-weighted, unadjusted Kaplan–Meier curves demonstrated progressively greater separation across phenotypes over the follow-up period of up to 240 months (Figure 4A). Estimated 10-year survival probabilities were 0.986 for no apparent risk, 0.892 for Stage A, 0.827 for Stage B, and 0.486 for Stage C. By the end of follow-up, survival probability had declined only modestly in the no-apparent-risk group (0.952), compared with more pronounced declines in Stage A (0.742) and Stage B (0.674). Stage C exhibited the steepest decline, with survival falling below 50% by month 115 and reaching 0.235 at the end of follow-up.

**Figure 4a.**
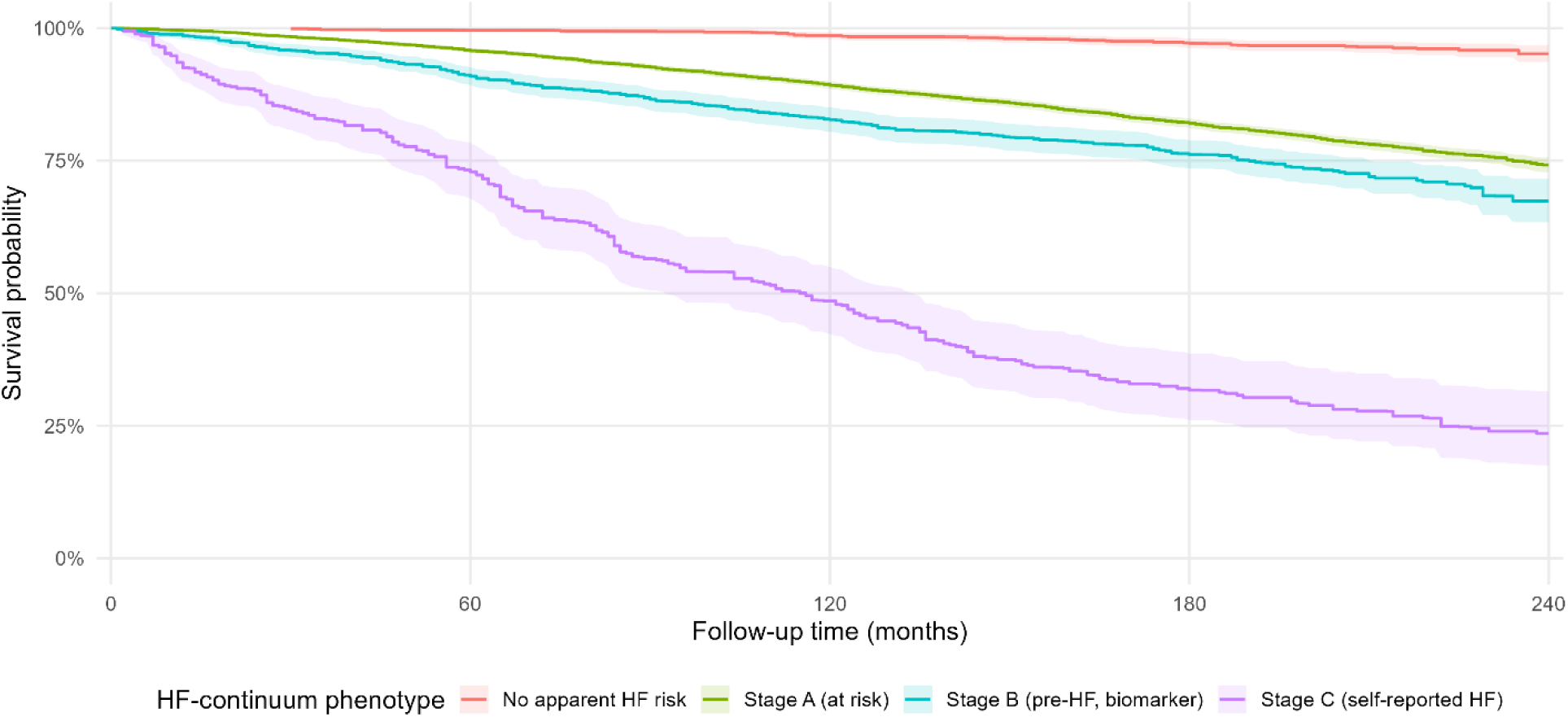
Survey-weighted unadjusted Kaplan-Meier curve.

**Figure 4b.**
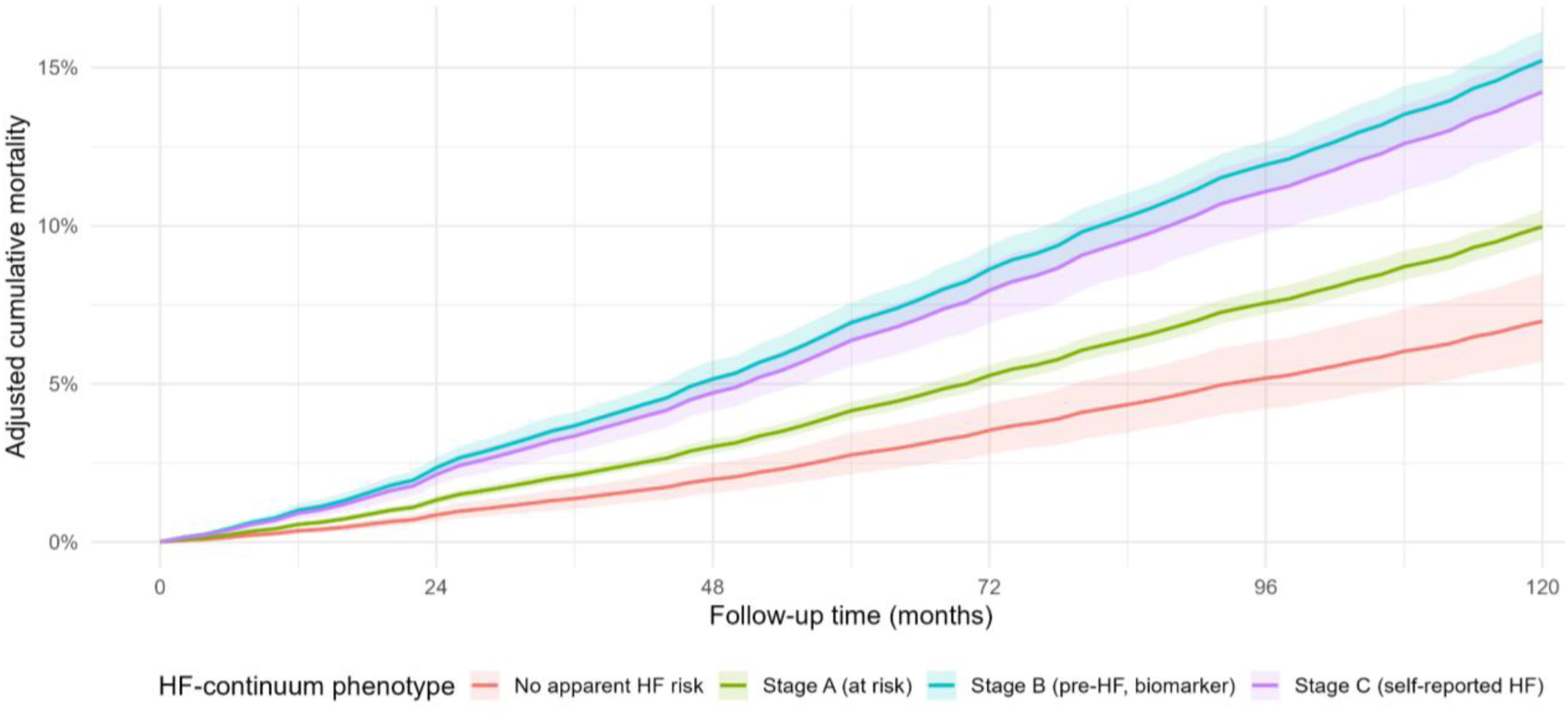
Adjusted cumulative mortality curve.

Hazard ratios (HRs) across HF phenotypes attenuated substantially with sequential covariate adjustment (Figure 5). In the unadjusted model (Model 0), HRs relative to the no-apparent-risk group were 6.54 (95% CI, 4.68–9.14) for Stage A, 8.94 (95% CI, 6.48– 12.34) for Stage B, and 37.43 (95% CI, 25.38–55.21) for Stage C. Adjustment for age, sex, and race and ethnicity (Model 1) reduced these estimates to 2.81, 5.60, and 6.68, respectively. Further adjustment for educational attainment, PIR, and insurance status (Model 2) yielded HRs of 2.18, 4.26, and 4.65, respectively. In the fully adjusted model (Model 3), which additionally accounted for hypertension, diabetes, obesity, CKD, smoking, and prior CHD/MI/stroke, the corresponding HRs were 1.58 (95% CI, 1.08–2.30) for Stage A, 2.88 (95% CI, 2.00–4.16) for Stage B, and 2.56 (95% CI, 1.61–4.09) for Stage C. Between Models 0 and 3, excess hazard (HR − 1) decreased by 95.7% for Stage C, 89.6% for Stage A, and 76.3% for Stage B. This attenuation likely reflects the older age and greater comorbidity burden of participants in more advanced phenotypes, as well as overlap between Model 3 covariates and the operational definition of Stage A.

**Figure 5.**
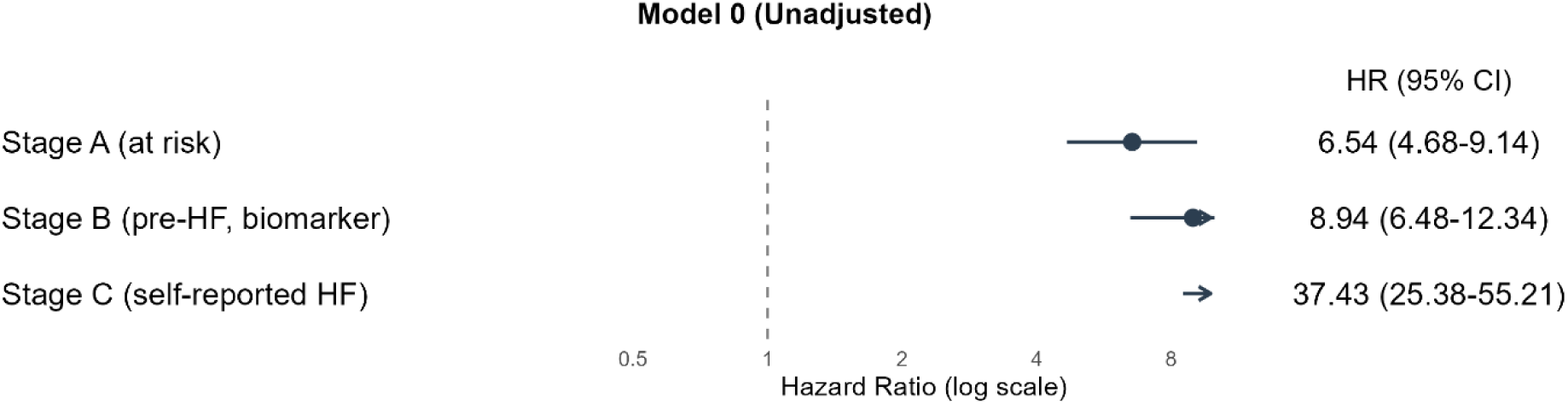
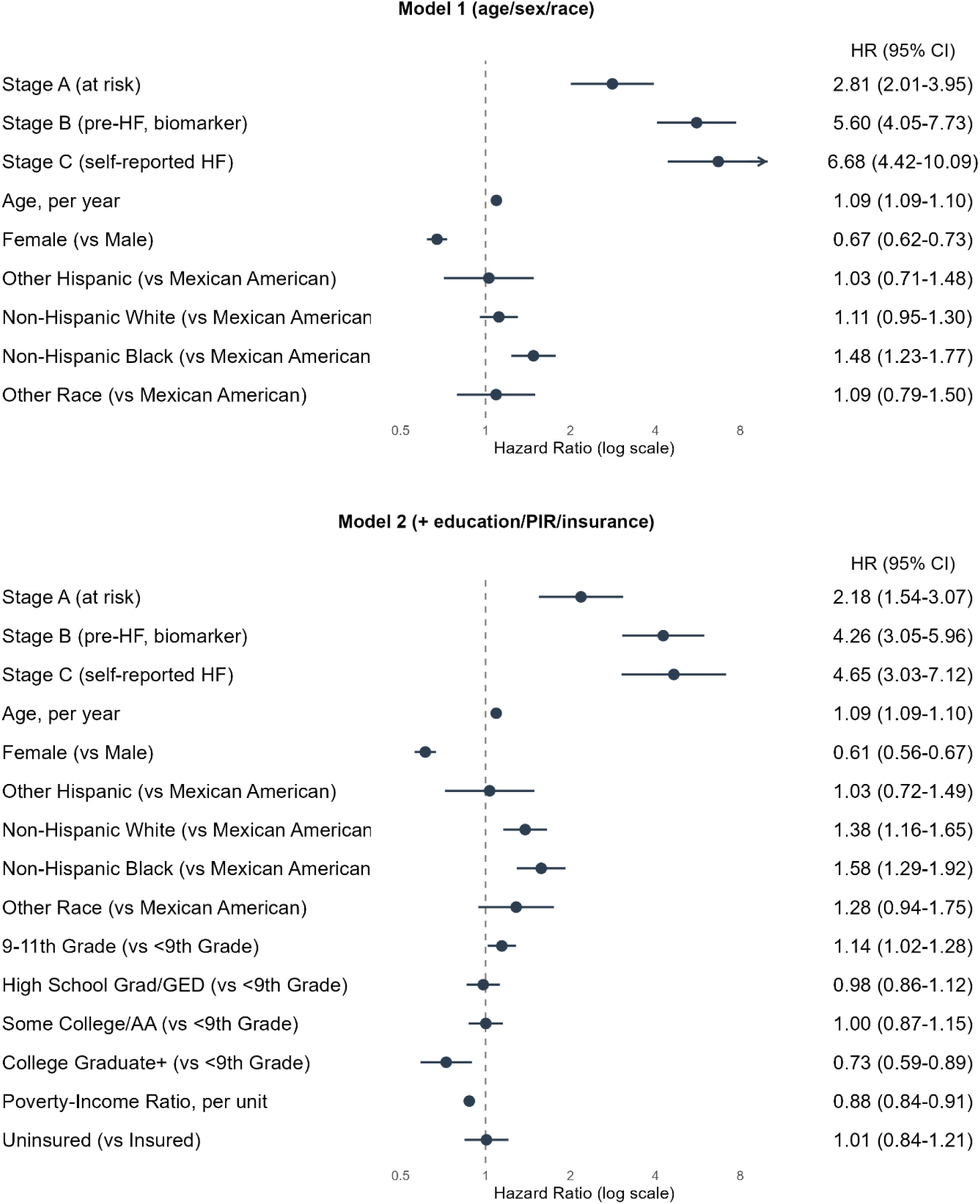
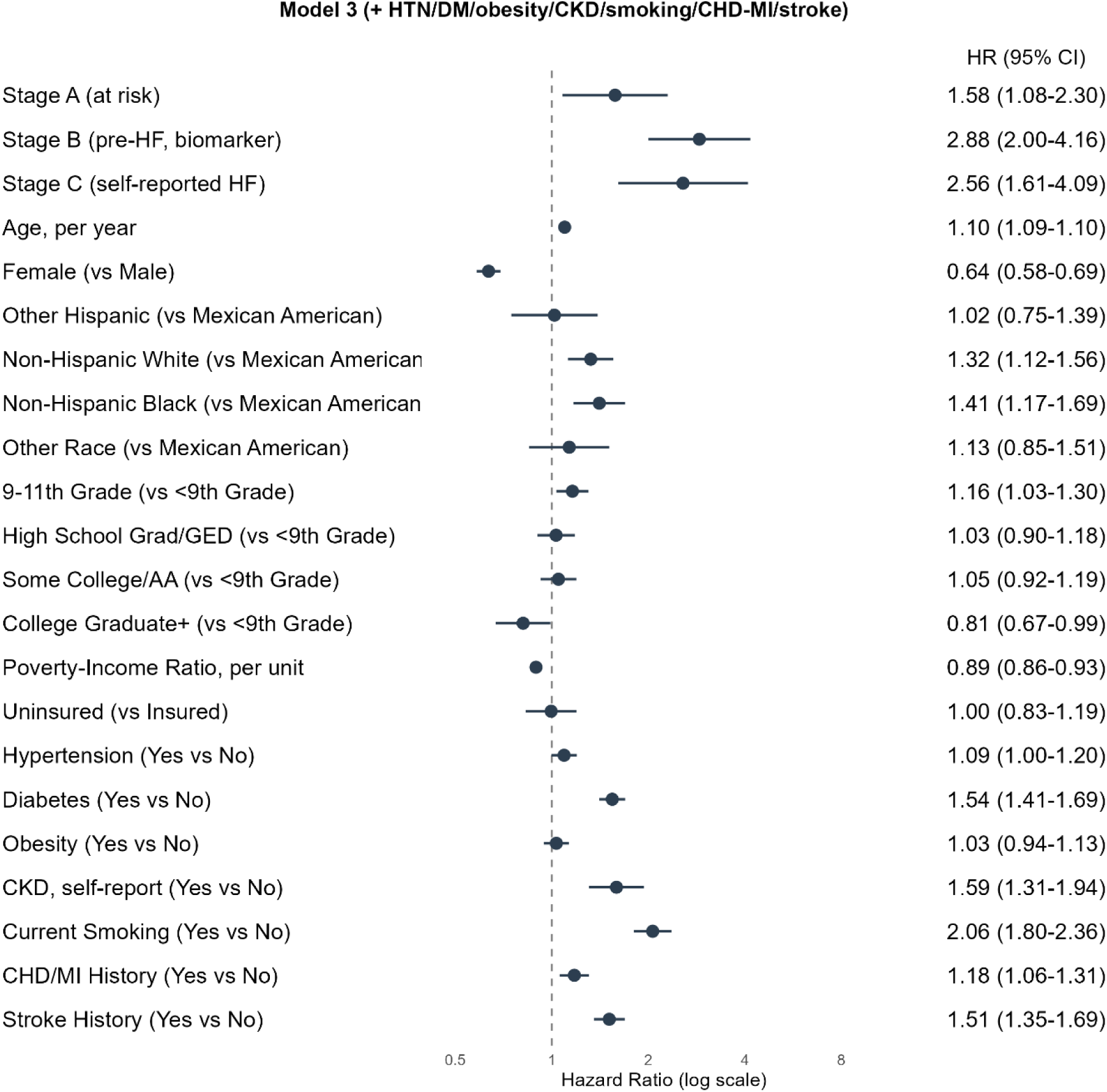
Hazard ratios for all-cause mortality across adjustment models. (a) Model 0: unadjusted; (b) Model 1: adjusted for age, sex, and race; (c) Model 2: additionally adjusted for education, PIR, and insurance; (d) Model 3: fully adjusted for multimorbidity and other covariates.

Among the covariates in Model 3, age (HR, 1.10 per year) and current smoking (HR, 2.06; 95% CI, 1.80–2.36) exhibited the largest associations with mortality, followed by CKD (HR, 1.59), diabetes (HR, 1.54), prior stroke (HR, 1.51), and prior CHD/MI (HR, 1.18). Hypertension and obesity were not significantly associated with mortality after adjustment. Female sex was associated with lower mortality (HR, 0.64; 95% CI, 0.58–0.69). Compared with Mexican American participants, mortality was higher among non-Hispanic Black (HR, 1.41) and non-Hispanic White participants (HR, 1.32). Higher PIR was associated with lower mortality (HR, 0.89 per unit; 95% CI, 0.86–0.93), while educational attainment exhibited a modest gradient. Insurance status was not significantly associated with mortality.

Because the preceding HRs were each estimated relative to the no-apparent-risk group, Stages A, B, and C were additionally compared directly to characterize differences in mortality risk across the more advanced phenotypes. Using Stage A as the reference, both Stage B (HR, 1.83; 95% CI, 1.62–2.07) and Stage C (HR, 1.63; 95% CI, 1.35–1.97) were associated with significantly higher mortality (both p<0.001). Because Stages A and B were both defined among participants without self-reported HF, this comparison provides an assessment of the additional prognostic information associated with biomarker-defined Stage B relative to Stage A, independent of classification into the self-report-dependent Stage C category. Conversely, when Stage B was used as the reference, the comparison with Stage C showed no statistically significant difference in mortality (HR, 0.89; 95% CI, 0.71–1.11; p=0.305) (Table 3). Notably, the point estimate for Stage B exceeded that for Stage C, indicating that the adjusted mortality pattern did not follow a strictly monotonic progression from no apparent risk through Stages A, B, and C.

**Table 3.** Pairwise comparison of hazard ratio among HF phenotypes.

| **Comparison** | **HR (95% CI)** | **p-value** |
|---|---|---|
| **Versus No Apparent HF Risk** |  |  |
| Stage A vs No apparent HF risk | 1.58 (1.08–2.30) | 0.018 |
| Stage B vs No apparent HF risk | 2.88 (2.00–4.16) | <0.001 |
| Stage C vs No apparent HF risk | 2.56 (1.61–4.09) | <0.001 |
| **Pairwise Comparisons** |  |  |
| Stage B vs Stage A | 1.83 (1.62–2.07) | <0.001 |
| Stage C vs Stage A | 1.63 (1.35–1.97) | <0.001 |
| Stage C vs Stage B | 0.89 (0.71–1.11) | 0.305 |

Standardized adjusted survival estimates, accounting for differences in covariate distributions across phenotypes, demonstrated a similar pattern, with the Stage B and Stage C survival curves converging. Estimated 10-year survival probabilities were 0.930 for no apparent risk, 0.900 for Stage A, 0.848 for Stage B, and 0.858 for Stage C, corresponding to adjusted 10-year mortality risks of 7.0%, 10.0%, 15.2%, and 14.2%, respectively. The corresponding 10-year restricted mean survival time (RMST) deficits relative to the no-apparent-risk group were 1.66 months for Stage A, 4.84 months for Stage B, and 4.21 months for Stage C (Figure 4B, Table 2). The observed 10-year mortality risk in the no-apparent-risk group was lower because standardization applies this phenotype classification across the covariate distribution of the entire analytic cohort, including participants with risk factors that preclude classification into this group. Thus, these estimates represent model-based predictions under hypothetical phenotype reassignment, with other covariates held constant, rather than mortality risks observed in the corresponding phenotype groups. Their reliability also depends on adequate covariate overlap between the phenotypes and the reference population. Using age, the covariate most likely to contribute to extrapolation, as an indicator of overlap, 68.9% of Stage A, 70.4% of Stage B, and only 30.3% of Stage C participants fell within the age range observed in the no-apparent-risk group. Accordingly, standardized estimates for Stage C are more heavily dependent on extrapolation beyond the observed reference-group age distribution.

### Incremental discriminative value of the Pre-HF biomarker phenotype

We evaluated whether incorporating the biomarker-defined stage improved prognostic performance beyond a conventional phenotype model. A reference model, in which participants originally classified as Stage B were reassigned to Stage A or no apparent risk according to their remaining classification criteria (75.7% and 24.3%, respectively), was compared with the corresponding model incorporating Stage B as a distinct category among all 11,477 participants (Table 4). The C-statistic increased marginally from 0.868 (95% CI, 0.863–0.873) to 0.870 (95% CI, 0.864–0.875), representing a statistically significant but clinically negligible improvement (ΔC-statistic, 0.002; 95% CI, 0.001– 0.002). The calibration slope remained similarly stable (0.965 vs. 0.969). The integrated discrimination improvement (IDI) was essentially null (0.0002; 95% CI, −0.0004 to 0.0008), whereas the continuous net reclassification improvement (NRI) was comparatively larger (0.219; 95% CI, 0.058–0.381). This discrepancy suggests that the NRI may be more sensitive to shifts in predicted risk than to meaningful improvements in overall discrimination. Collectively, these findings indicate that the biomarker-defined stage carries prognostic information but provides little incremental improvement in population-level risk discrimination beyond conventional predictors.

**Table 4.** Incremental prognostic value of biomarker-defined Stage B/pre-HF.

| **Metric** | **Reference Model<sup>†</sup>** | **Model + Stage B** |
|---|---|---|
| C-statistic (95% CI) | 0.868 (0.863-0.873) | 0.870 (0.864-0.875) |
| Δ C-statistic (95% CI) | Reference | 0.002 (0.001-0.002) |
| Calibration slope (95% CI) | 0.965 (0.955-0.975) | 0.969 (0.958-0.981) |
| IDI<sup>‡</sup> (95% CI) | — | 0.0002 (-0.0004-0.0008) |
| NRI<sup>§</sup> (95% CI) | — | 0.219 (0.058-0.381) |

<sup>†</sup> Reference model: Model 3 covariates plus a 3-level phenotype in which participants originally meeting Stage B criteria were reassigned to Stage A or no apparent risk according to the remaining classification criteria. Both models were fit on the identical N=11,477 participants;
<sup>‡</sup> IDI: Integrated Discrimination Improvement;
<sup>§</sup> NRI: Net Reclassification Improvement.

### Sensitivity analysis

The association between Stage B and all-cause mortality was directionally consistent and similar in magnitude across six alternative analytical specifications (Table 5), compared with the primary analysis (Model 3; pooled multiple-imputation estimate: HR, 2.88; 95% CI, 2.00–4.16; N = 11,477).

**Table 5.**
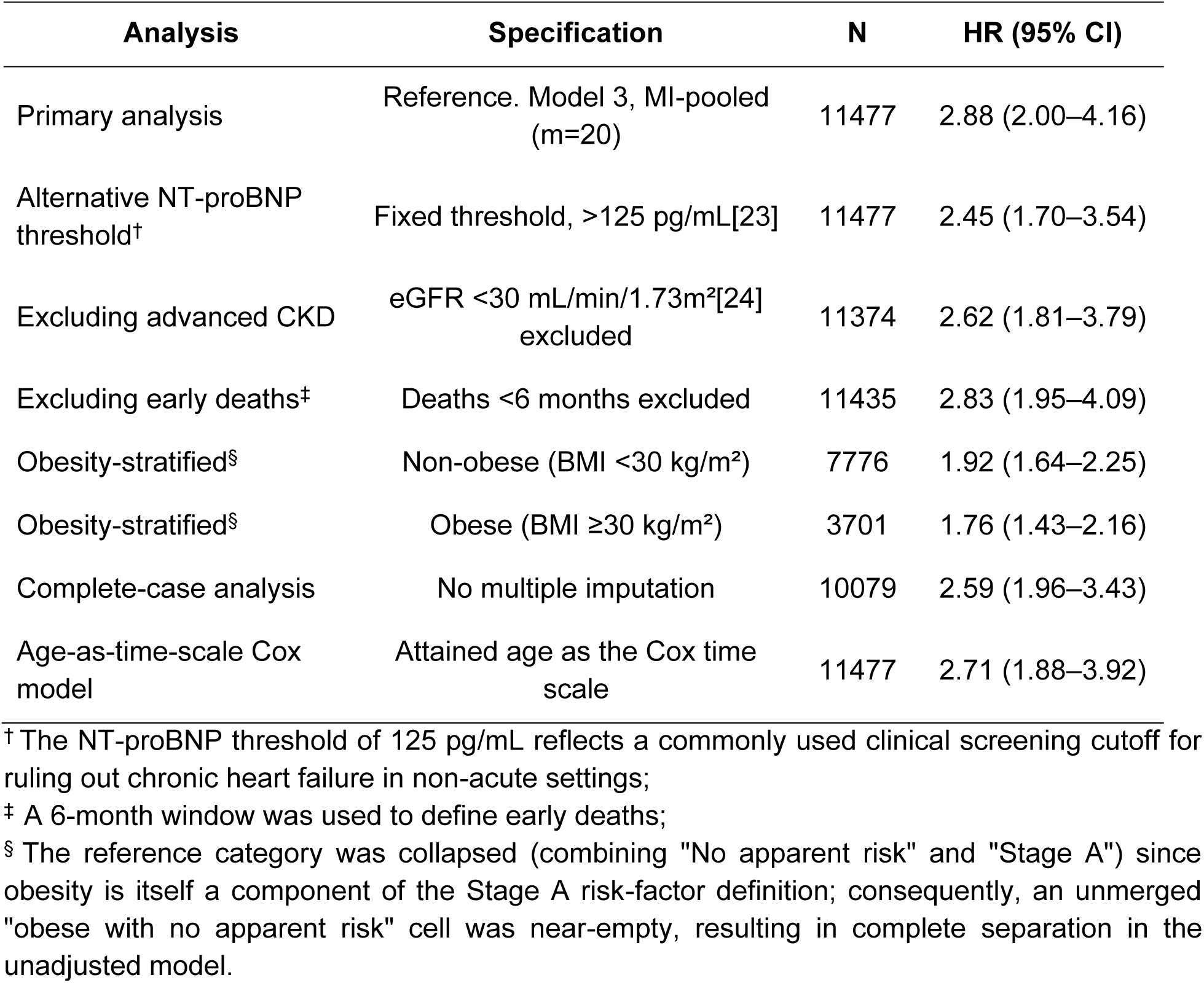
Sensitivity Analyses of Stage B/Pre-HF Mortality Association.

| Analysis | Specification | N | HR (95% CI) |
|---|---|---|---|
| Primary analysis | Reference. Model 3, MI-pooled (m=20) | 11477 | 2.88 (2.00–4.16) |
| Alternative NT-proBNP threshold<sup>†</sup> | Fixed threshold, >125 pg/mL[23] | 11477 | 2.45 (1.70–3.54) |
| Excluding advanced CKD | eGFR <30 mL/min/1.73m²[24] excluded | 11374 | 2.62 (1.81–3.79) |
| Excluding early deaths<sup>‡</sup> | Deaths <6 months excluded | 11435 | 2.83 (1.95–4.09) |
| Obesity-stratified<sup>§</sup> | Non-obese (BMI <30 kg/m²) | 7776 | 1.92 (1.64–2.25) |
| Obesity-stratified<sup>§</sup> | Obese (BMI ≥30 kg/m²) | 3701 | 1.76 (1.43–2.16) |
| Complete-case analysis | No multiple imputation | 10079 | 2.59 (1.96–3.43) |
| Age-as-time-scale Cox model | Attained age as the Cox time scale | 11477 | 2.71 (1.88–3.92) |

<sup>†</sup> The NT-proBNP threshold of 125 pg/mL reflects a commonly used clinical screening cutoff for ruling out chronic heart failure in non-acute settings;
<sup>‡</sup> A 6-month window was used to define early deaths;
<sup>§</sup> The reference category was collapsed (combining "No apparent risk" and "Stage A") since obesity is itself a component of the Stage A risk-factor definition; consequently, an unmerged "obese with no apparent risk" cell was near-empty, resulting in complete separation in the unadjusted model.

Replacing the age-and sex-specific 90th-percentile definition with a fixed clinical NT-proBNP threshold (>125 pg/mL) yielded a modestly attenuated but statistically significant association (HR, 2.45; 95% CI, 1.70–3.54). Exclusion of participants with advanced CKD (eGFR <30 mL/min/1.73 m²; N = 11,374) and exclusion of deaths occurring within the first 6 months of follow-up (N = 11,435) yielded estimates comparable to the primary analysis (HR, 2.62; 95% CI, 1.81–3.79, and HR, 2.83; 95% CI, 1.95–4.09, respectively). In analyses stratified by obesity status and using a combined reference category comprising no apparent risk and Stage A, the association was modestly stronger among non-obese participants (BMI <30 kg/m²; HR, 1.92; 95% CI, 1.64–2.25; N = 7,776) than among obese participants (BMI ≥30 kg/m²; HR, 1.76; 95% CI, 1.43–2.16; N = 3,701), although the confidence intervals overlapped. Complete-case analysis without multiple imputation (N = 10,079) and an alternative Cox model using age as the underlying time scale also produced estimates consistent with the primary findings (HR, 2.59; 95% CI, 1.96–3.43, and HR, 2.71; 95% CI, 1.88–3.92, respectively).

## DISCUSSION

### Feasibility of a survey-based pre-HF phenotype

Mortality risk increased across the phenotypic continuum, from no apparent risk through Stage A to Stages B and C. After full covariate adjustment, both Stage B and Stage C remained associated with significantly higher mortality than Stage A; however, their relative ordering was not statistically established, with the point estimate for Stage B modestly exceeding that for Stage C. Thus, the observed risk gradient distinguished no apparent risk, Stage A, and the two more advanced phenotypes, without demonstrating a strictly monotonic progression between Stage B and Stage C. These findings suggest that the updated consensus criteria for pre-HF, which incorporate biomarker elevation as an alternative to imaging-based evidence of cardiac abnormality, can be approximated using routinely collected variables from a national health survey.[25] This approach may have implications for population-level surveillance and epidemiologic research, particularly in settings where echocardiographic assessment is not routinely available.[26, 27]

### Stage B and covariate adjustment

The associations of both Stage B and Stage C with mortality attenuated progressively with sequential covariate adjustment. In the unadjusted model, the HR for Stage C substantially exceeded that for Stage B, consistent with Stage C representing clinically overt disease. However, excess hazard decreased by 95.7% for Stage C and 76.3% for Stage B between the unadjusted and fully adjusted models, resulting in convergence of their HRs. This differential attenuation likely reflects differences in age and comorbidity burden across phenotypes, particularly the considerably older age distribution of Stage C participants, rather than indicating that the Stage B association is less attributable to conventional risk factors. Furthermore, because several covariates in Model 3 overlap with the operational definition of Stage A, this pattern characterizes changes in associations across the specified adjustment sequence and does not establish any particular underlying mechanism. Nevertheless, Stage B remained associated with significantly higher mortality than no apparent risk after full adjustment, supporting the prognostic relevance of the biomarker-defined phenotype.

These findings also help reconcile an apparent discrepancy between the adjusted mortality associations and the incremental prognostic performance of the phenotype. Although Stage B retained a significantly elevated HR in the fully adjusted model, its incorporation into the conventional model yielded only a negligible improvement in the C-statistic (0.868 to 0.870) and an IDI estimate that was not distinguishable from zero. The continuous NRI was nominally significant (0.219; 95% CI, 0.058–0.381); however, this metric can be sensitive to relatively small changes in predicted risk, even in the absence of meaningful improvements in discrimination. These measures address different aspects of prognostic performance: an HR quantifies the relative mortality hazard associated with a phenotype, whereas discrimination and reclassification metrics assess whether incorporating that phenotype improves risk ranking or classification across individuals in a population. The latter objective becomes inherently more difficult when a strong predictor, such as age, is already included in the model.[28] Collectively, these findings support the prognostic relevance of biomarker-defined Stage B beyond conventional predictors but do not establish this particular operational phenotype as a validated clinical entity. An important related question—whether categorical staging of NT-proBNP provides additional prognostic information beyond its use as a continuous biomarker— was not directly evaluated and warrants further investigation.

### Robustness across analytical specifications

The association between Stage B and all-cause mortality was directionally consistent and of broadly similar magnitude across alternative analytical specifications, including use of a fixed NT-proBNP threshold, exclusion of participants with advanced CKD, exclusion of early deaths, complete-case analysis, and use of age as the underlying time scale in Cox models. One potential exception emerged in the obesity-stratified analysis, in which the association appeared modestly weaker among participants with obesity. However, the confidence intervals overlapped substantially, and no formal test for interaction was performed. A plausible explanation is the well-documented inverse relationship between obesity and circulating natriuretic peptide concentrations, reported since at least the mid-2000s and potentially attributable to reduced cardiac production, altered peptide clearance, or both.[29, 30] Consequently, applying a population-based percentile threshold may result in lower sensitivity for identifying biomarker-defined Stage B among individuals with obesity. This possibility warrants further investigation, particularly through formal interaction testing and evaluation of obesity-specific biomarker thresholds in future studies.

### Limitations and future directions

There are several limitations. First, the phenotype is a proxy-based approximation of the 2026 Universal Definition, with important limitations in stage assignment. Elevated NT-proBNP alone cannot establish pre-HF, as elevations may reflect noncardiac conditions.

Similarly, absence of self-reported HF does not exclude unrecognized symptoms, potentially misclassifying symptomatic individuals as Stage B, while self-reported HF cannot distinguish Stage C from Stage D. Participants meeting none of the Stage A criteria are better characterized as having no measured HF risk factors, as structural cardiac disease could not be assessed.[31] Moreover, defining Stage B by the 90^th^ percentile inherently influenced its prevalence. The absence of echocardiographic data in NHANES,[32] precluded classification of Stage D, ejection fraction phenotypes, worsening HF, and remission or recovery. These limitations underscore the challenges of translating multimodal staging frameworks into survey-based phenotypes and the need for validation against imaging-confirmed Stage B.[33] Second, the temporal scope of the biomarker data limits generalizability to contemporary populations. NT-proBNP was measured only in stored serum samples from the 1999–2004 NHANES cycles,[16] during which background risk-factor prevalence, treatment patterns, and laboratory assay practices may have differed substantially from those of current populations. Third, phenotype assignment was based on a single cross-sectional examination, capturing each participant’s classification at one time point rather than characterizing longitudinal transitions. Cohorts with repeated biomarker, clinical, and imaging assessments are needed to determine the frequency, timing, and predictors of transitions between HF stages.[34] Fourth, residual missing data were addressed using deterministic within-stratum imputation, and the 90th-percentile NT-proBNP threshold was statistically derived rather than clinically validated. Although sensitivity analyses using an alternative threshold and excluding advanced CKD supported the robustness of findings, they cannot exclude age-or kidney-related influences on NT-proBNP. Fifth, all-cause mortality cannot establish whether advanced phenotypes reflect cardiac disease progression or cardiovascular-specific risk. A related NHANES analysis using risk factor-based preclinical HF definitions reported stronger associations with cardiovascular than all-cause mortality, suggesting that our endpoint may underestimate cardiovascular-specific associations.[13] However, cause-specific analyses require validated cardiovascular death classification and appropriate competing-risk methods, as noncardiovascular death precludes subsequent cardiovascular death.[35]

## CONCLUSION

This study examined whether the 2026 Universal Definition of Heart Failure could be operationalized as a computable, biomarker-informed phenotype using a nationally representative U.S. health survey and whether the resulting classification provided meaningful prognostic information. Using self-reported HF status, measured NT-proBNP, and conventional cardiometabolic risk factors from NHANES, we constructed a four-level HF-continuum phenotype that stratified long-term all-cause mortality risk. The highest mortality risks were observed in Stages B and C, which exhibited HRs of similar magnitude after full covariate adjustment. Stage B remained independently associated with mortality after adjustment, and its association was not fully attenuated by the covariates that accounted for much of the excess mortality associated with Stage C. However, incorporating Stage B into a conventional risk model yielded only marginal improvements in discrimination, highlighting the distinction between an independent prognostic association and incremental improvement in population-level risk stratification.

## Data Availability

All data produced are available online at https://www.cdc.gov/nchs/nhanes/

## DECLARATIONS

## Ethics approval and consent to participate

NHANES was approved by the National Center for Health Statistics Research Ethics Review Board, and informed consent was obtained from all participants; the relevant ethical approval protocol for the 1999–2004 survey cycles used in this analysis was Protocol #98-12.

## Consent for publication

This study used data from the NHANES database, a publicly available, de-identified database. As the study involved secondary analysis of existing de-identified data and did not involve direct interaction with human participants or access to identifiable private information, informed consent and Institutional Review Board (IRB) approval were waived.

## Availability of data and materials

All NHANES data used in this analysis are publicly available from the National Center for Health Statistics (https://www.cdc.gov/nchs/nhanes/). The code for cohort construction and all analyses is available at: https://github.com/hxcconnie-star/biomarker_phenotype_hf.

## Authors’ contributions

JY developed the study. HC and JY contributed to the data analyses, interpretation of the results, and the writing of the manuscript. All authors approved the final version of the manuscript.

## Acknowledgements

None.

## Supplementary

**Table S1.**
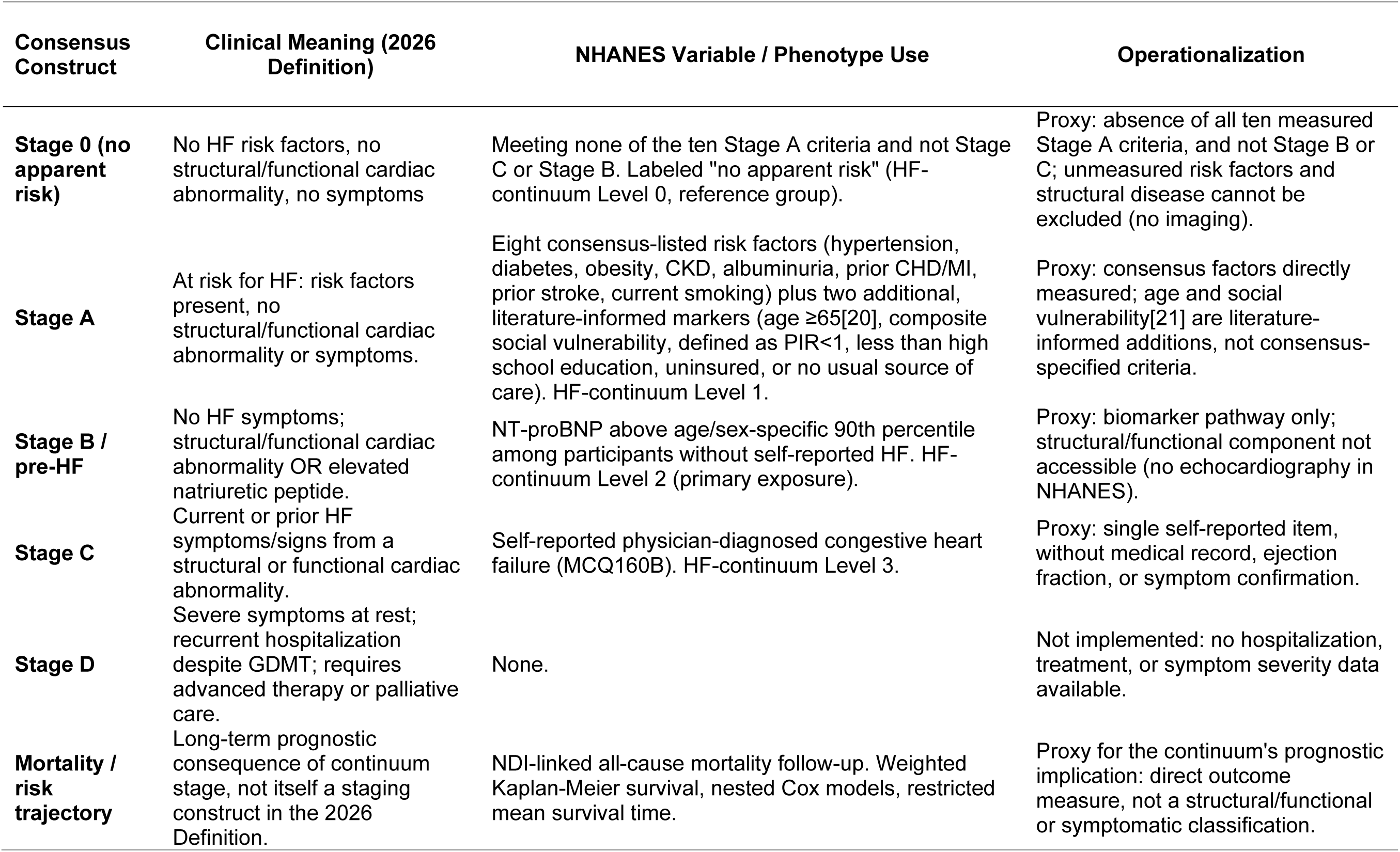
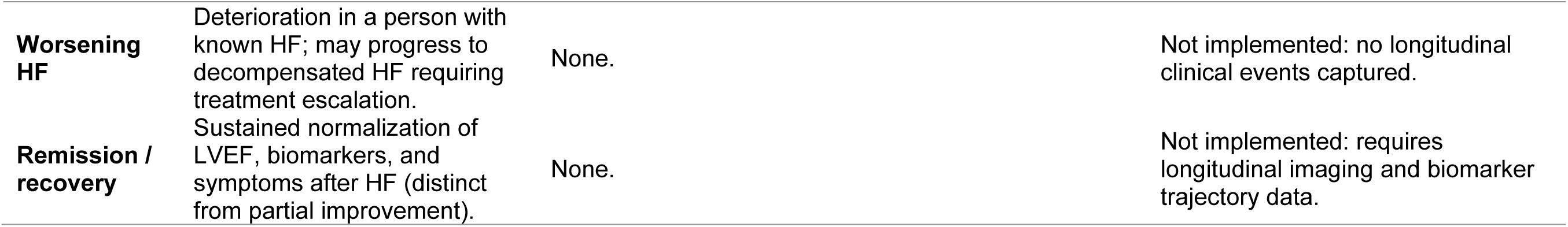
Mapping of 2026 consensus constructs to NHANES variables.

| Consensus Construct | Clinical Meaning (2026 Definition) | NHANES Variable / Phenotype Use | Operationalization |
|---|---|---|---|
| **Stage 0 (no apparent risk)** | No HF risk factors, no structural/functional cardiac abnormality, no symptoms | Meeting none of the ten Stage A criteria and not Stage C or Stage B. Labeled "no apparent risk" (HF-continuum Level 0, reference group). | Proxy: absence of all ten measured Stage A criteria, and not Stage B or C; unmeasured risk factors and structural disease cannot be excluded (no imaging). |
| **Stage A** | At risk for HF: risk factors present, no structural/functional cardiac abnormality or symptoms. | Eight consensus-listed risk factors (hypertension, diabetes, obesity, CKD, albuminuria, prior CHD/MI, prior stroke, current smoking) plus two additional, literature-informed markers (age ≥65[20], composite social vulnerability, defined as PIR<1, less than high school education, uninsured, or no usual source of care). HF-continuum Level 1. | Proxy: consensus factors directly measured; age and social vulnerability[21] are literature-informed additions, not consensus-specified criteria. |
| **Stage B / pre-HF** | No HF symptoms; structural/functional cardiac abnormality OR elevated natriuretic peptide. | NT-proBNP above age/sex-specific 90th percentile among participants without self-reported HF. HF-continuum Level 2 (primary exposure). | Proxy: biomarker pathway only; structural/functional component not accessible (no echocardiography in NHANES). |
| **Stage C** | Current or prior HF symptoms/signs from a structural or functional cardiac abnormality. | Self-reported physician-diagnosed congestive heart failure (MCQ160B). HF-continuum Level 3. | Proxy: single self-reported item, without medical record, ejection fraction, or symptom confirmation. |
| **Stage D** | Severe symptoms at rest; recurrent hospitalization despite GDMT; requires advanced therapy or palliative care. | None. | Not implemented: no hospitalization, treatment, or symptom severity data available. |
| **Mortality / risk trajectory** | Long-term prognostic consequence of continuum stage, not itself a staging construct in the 2026 Definition. | NDI-linked all-cause mortality follow-up. Weighted Kaplan-Meier survival, nested Cox models, restricted mean survival time. | Proxy for the continuum's prognostic implication: direct outcome measure, not a structural/functional or symptomatic classification. |
| **Worsening HF** | Deterioration in a person with known HF; may progress to decompensated HF requiring treatment escalation. | None. | Not implemented: no longitudinal clinical events captured. |
| **Remission / recovery** | Sustained normalization of LVEF, biomarkers, and symptoms after HF (distinct from partial improvement). | None. | Not implemented: requires longitudinal imaging and biomarker trajectory data. |

**Table S2.**
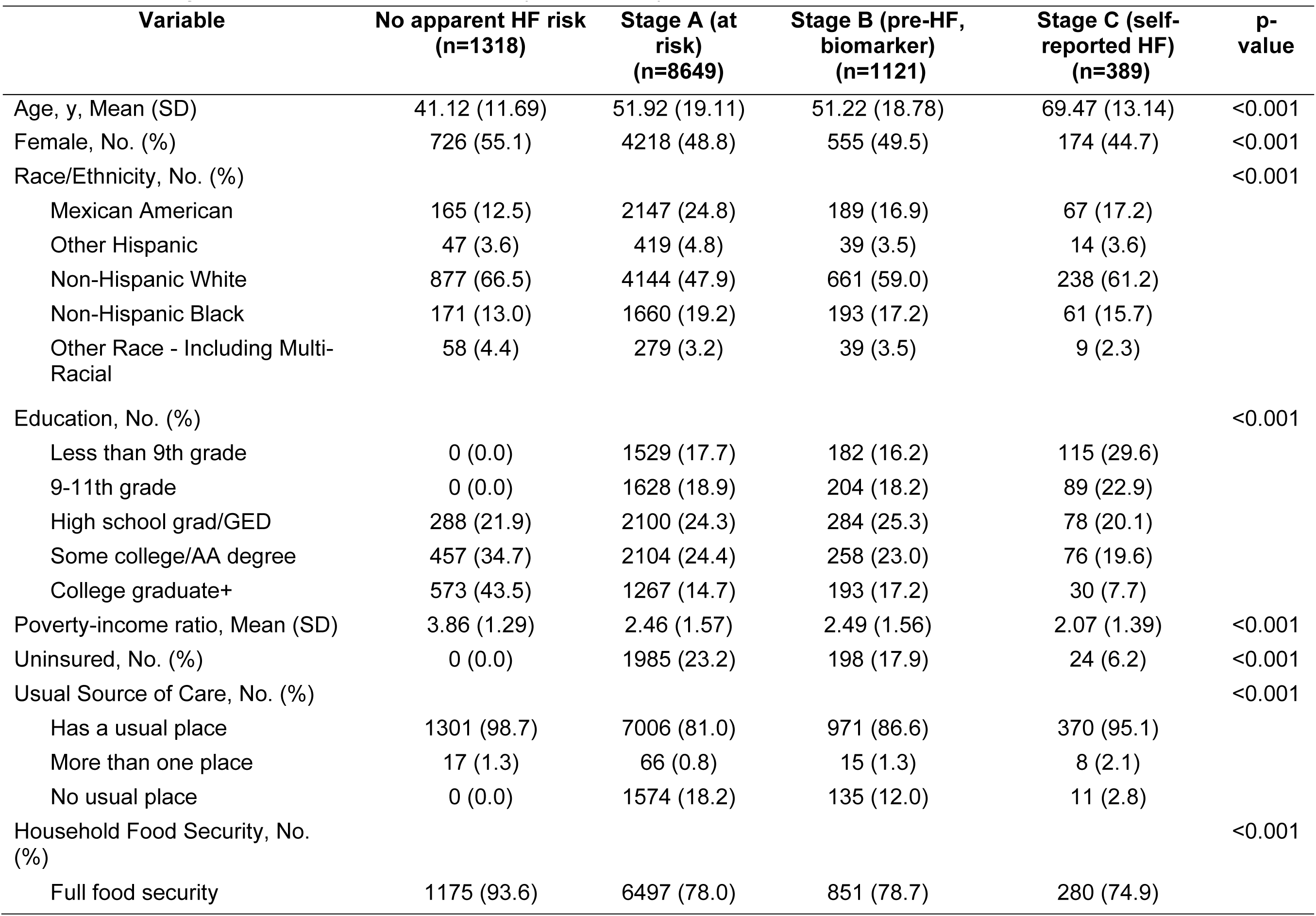
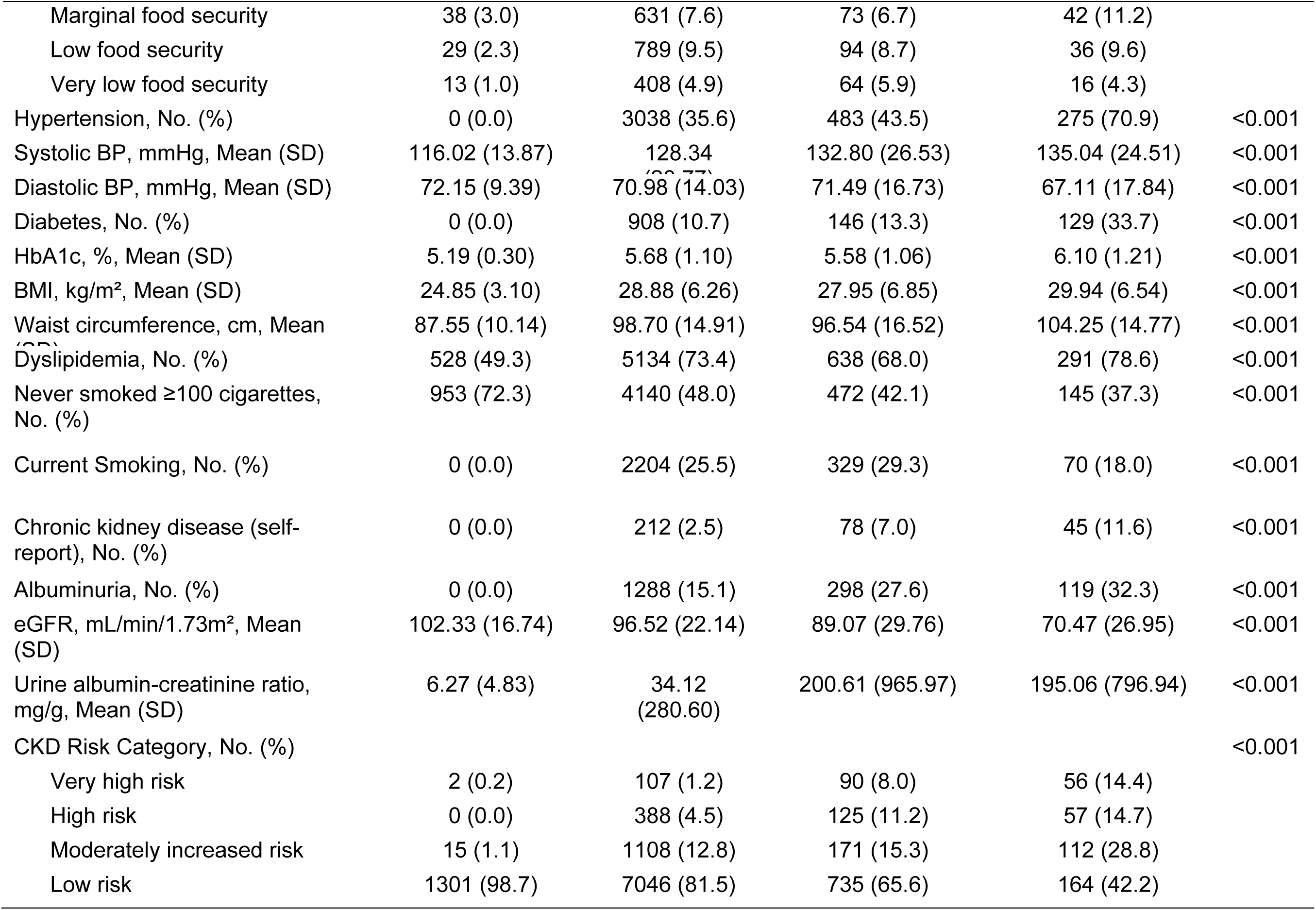
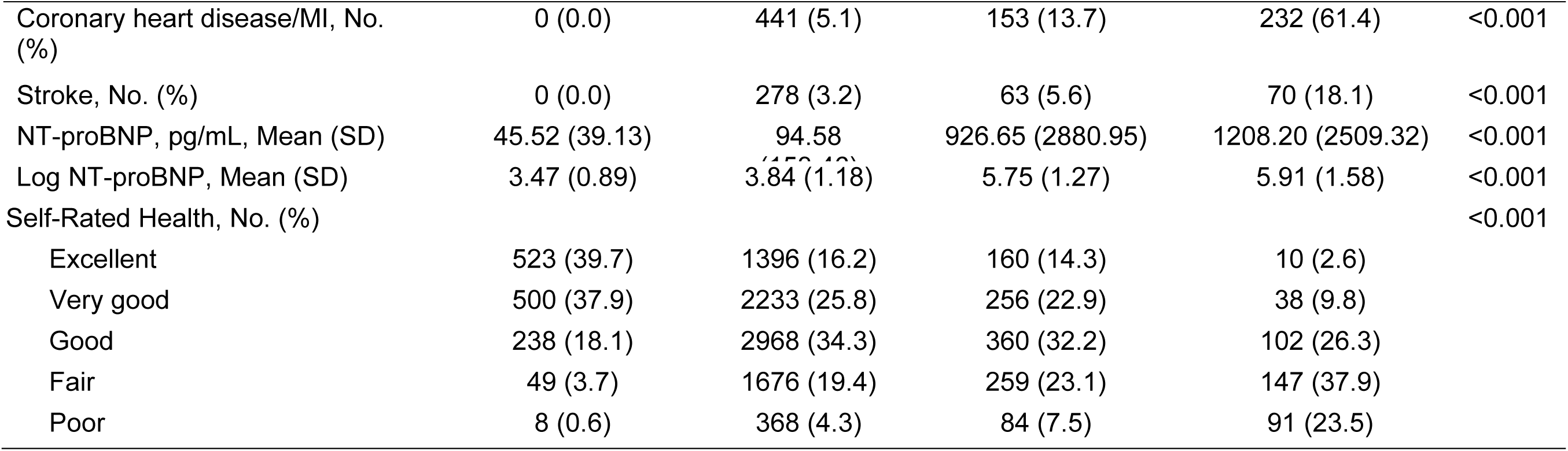
Unweighted baseline characteristics by HF phenotype.

